# Facets of language performance in early-onset and late-onset Alzheimer’s disease dementia

**DOI:** 10.1101/2025.06.13.25329521

**Authors:** Jeanne Gallée, Laura E. Gibbons, Seo-Eun Choi, Michael Lee, Phoebe Scollard, Emily H. Trittschuh, Jesse Mez, Andrew J. Saykin, Nancy S. Foldi, Shubhabrata Mukherjee, Paul K. Crane

**Affiliations:** Center for Psychometric Analyses of Aging and Neurodegeneration, Department of Medicine, University of Washington, Seattle, WA 98195, USA; Alzheimer’s Disease Research Center, University of Washington, Seattle WA 98195, USA; University of Bordeaux, Inserm, Bordeaux Population Health Research Center (UMR 1219), 33000 Bordeaux, France; Department of Psychiatry and Behavior Sciences, University of Washington School of Medicine, Seattle, WA 98195, USA; VA Puget Sound Health Care System, GRECC, Seattle, WA 98108, USA; Department of Neurology, Boston University Chobanian & Avedisian School of Medicine, Boston, MA 02118, USA; Department of Radiology and Imaging Sciences, Indiana University School of Medicine, Indianapolis, IN 46202, USA; Indiana Alzheimer’s Disease Research Center, Indiana University School of Medicine, Indianapolis, IN 46202, USA; Stark Neurosciences Research Institute, Indiana University School of Medicine, Indianapolis, IN 46202, USA; Department of Radiology, Brain Health Imaging Institute, Weill Cornell Medicine, New York, New York, 10065 USA

**Keywords:** language, Alzheimer’s disease dementia, early-onset, late-onset, non-amnestic

## Abstract

**BACKGROUND:** Early-onset Alzheimer’s disease dementia (EOAD) is characterized by more pronounced cognitive decline than late-onset AD dementia (LOAD). Characteristic performance in spoken language remains undefined.

**METHOD:** A cross-sectional analysis of 1,189 people with EOAD and 4,646 with LOAD from the National Alzheimer’s Coordinating Center (NACC).

**RESULT:** Based on data from their first NACC visit with AD, there was considerable heterogeneity in language performance across people with EOAD and LOAD. The distribution of naming ability was similar across these groups. On average, people with LOAD had better performance than people with EOAD for category fluency, letter fluency, and spoken lexical retrieval, and had lower Clinical Dementia Rating (CDR®) language scores, though there was considerable overlap in all of the distributions for people with EOAD and people with LOAD.

**DISCUSSION:** At diagnosis, EOAD and LOAD language profiles are distinct. There is substantial variability in both groups in multiple aspects of language.

## 1.0 INTRODUCTION

Alzheimer’s disease (AD) pathology is the most common cause of dementia, accounting for over half of cases worldwide [1]. AD dementia is a clinical syndrome characterized by multi-domain cognitive decline, including memory, accompanied by acquired functional impairment in a major life category [2–4]. Two subtypes are identified by age at diagnosis: early-onset (EOAD) and late-onset AD dementia (LOAD). While EOAD and LOAD are defined by objective impairments in the memory domain, changes other cognitive domains are frequently present at diagnosis. The clinical presentation of LOAD is increasingly recognized as heterogeneous [5,6], with emerging evidence for distinct cognitive subgroups demarcated by relative impairments across domains [7–9]. Similar heterogeneity at the time of dementia diagnosis is also appreciated in people with EOAD [5,10,11].

The impact of AD dementia on the language domain has received growing attention. Subtle changes to language, such as to lexical retrieval (e.g., naming [6] and verbal fluency [12]), linguistic complexity (e.g., word frequency and grammatical structure [13]), and linguistic understanding (e.g., written or auditory comprehension [14]), may emerge early in AD dementia. Spoken language performance—evaluated through measures of naming, verbal fluency, and spontaneous speech—is indicative of cognitive decline and clinically differentiates between MCI and dementia [12,15–17].

Characterizations of language performance differences between EOAD and LOAD remain mixed. Certain studies report greater impairment for people with EOAD for elements of verbal learning [18], comprehension [19], writing [20], and verbal letter fluency [21], with more rapid decline on average [10,11,19,20,22]. Other investigations support that people with LOAD present with greater decline across multiple language subdomains [6,14], including semantics [23], confrontation naming ability [19,21,22,24], and verbal fluency [25]. Others report inconsequential baseline differences in language performance on average between people with EOAD and people with LOAD [11,26]. These discrepancies may stem from methodological challenges, such as limited sample sizes, single-site data, or the reliance on coarse screening tools such as the Mini-Mental State Examination (MMSE [27]) or the Montreal Cognitive Assessment (MoCA [28]), which capture language performance to a limited extent.

Consequently, there remain considerable gaps in scientific understanding of various aspects of language in AD dementia. This study addresses these gaps by characterizing facets of language performance in a large, well-phenotyped sample of people with EOAD or LOAD. We focus on granular elements of spoken lexical generation through core tasks of neuropsychological assessment (e.g., naming and verbal fluency). Spoken language is a central function of daily communication with direct implications for potential behavioral intervention in the absence of curative pharmacological treatment [29–35].

This study leverages the extraordinarily rich data resources of the National Alzheimer’s Coordinating Center (NACC) [36] dataset, representing a well-characterized and multicenter sample of people living with AD. The dataset is uniquely suited for this analysis due to its breadth, rigorous diagnostic procedures, standardized collection by trained clinicians, and its large representation of EOAD—a population often underrepresented in large-scale studies These strengths position the current work to provide novel insight into characteristic language performance in EOAD and LOAD, aligning with ongoing efforts to advance dementia subtyping and characterization.

## 2.0 MATERIALS & METHODS

### 2.1 Participants

Participant data was drawn from the NACC Uniform Data Set 3.0 Neuropsychological Battery (UDS3-NB [36]). There were 26,157 participants in UDS3-NB with any language items (66,112 records). Individuals who did not have AD as a primary or secondary etiology (variable: naccalzd) were excluded from the dataset. From the remaining 10,017 participants, 4,148 for whom the variable naccudsd was not “Dementia” were removed. For these 5,869 participants, only the record that corresponded to the first visit at which an AD diagnosis was formulated were kept. Finally, 34 participants with a global CDR® [37] score of 0 (a categorical score denoting normal functional performance across domains of memory, orientation, judgement and problem-solving, community affairs, home and hobbies, and personal care) were removed from the dataset. We categorized participants as EOAD if they were younger than 65 at the time of their first visit with a diagnosis of AD in UDS1, UDS2, or UDS3. Our classification resulted in a participant sample of 1,189 EOAD and 4,646 LOAD.

### 2.2 Harmonized Language Score

We implemented the harmonization workflow as stated in Mukherjee *et. al.* [38] to harmonize the language domain in NACC. Briefly, qualified neuropsychologists and behavioral neurologists categorized NACC test items (UDS1, UDS2, and UDS3) into memory, executive functioning, language, visuospatial, or none of these domains. The analytic team evaluated each NACC test item with the cognitive specialist panel to ensure administration and scoring are equivalent for anchor items. Items were treated as categorical and recoded to a maximum of 10 categories as needed. Overlapping test items across NACC and our item bank (data derived from additional previously harmonized and co-calibrated data sets) were treated as anchor items. Test items were indicators in a confirmatory factor analysis (CFA) model, with all anchor item parameters fixed and non-overlapping test items freely estimated. The CFA model was run on the most recent visit for each individual for a given data freeze (NACC freeze April 2024) to obtain item parameters (factor loadings and thresholds) for unique NACC items. These item parameters were applied to the longitudinal data set (e.g. all visits, not just the most recent visit) to obtain factor scores for the language domain (Supplementary Figure 1). The language items that were part of the tests implemented in UDS3 are in Table 2.

**Table 1.** NACC Participant Characteristics.

|  | AD in NACC before age 65 |  |  | p-value <sup>1</sup> |
| --- | --- | --- | --- | --- |
|  | No<br>(n=4646) | Yes<br>(n=1189) | Total<br>(n=5835) |  |
| Age | 78.1 (7.4) | 58.8 (4.9) | 74.2 (10.4) | < 0.001 |
| Female | 2,395 (51.5%) | 661 (55.6%) | 3,056 (52.4%) | 0.013 |
| Education |  |  |  | < 0.001 |
| <= High school | 1,094 (23.7%) | 278 (23.6%) | 1,372 (23.7%) |  |
| Some/all college | 1,843 (40.0%) | 582 (49.4%) | 2,425 (41.9%) |  |
| Post college | 1,676 (36.3%) | 317 (26.9%) | 1,993 (34.4%) |  |
| Race |  |  |  |  |
| American Indian or Alaska Native | 21 (0.5%) | 9 (0.8%) | 30 (0.5%) | 0.001 |
| Asian | 117 (2.5%) | 24 (2.0%) | 141 (2.4%) |  |
| Black or African American | 449 (9.8%) | 65 (5.5%) | 514 (8.9%) |  |
| Native Hawaiian or Other Pacific Islander | 5 (0.1%) | 5 (0.4%) | 10 (0.2%) |  |
| Other | 70 (1.5%) | 14 (1.2%) | 84 (1.5%) |  |
| White | 3,942 (85.6%) | 1,055 (90.0%) | 4,997 (86.5%) |  |
| Any *APOE* ε4 alleles | 2,515 (54.1%) | 711 (59.8%) | 3,226 (55.3%) | < 0.001 |
| CDR sum of boxes | 5.5 (3.2) | 5.7 (3.4) | 5.6 (3.2) | 0.201 |
| CDR language |  |  |  | < 0.001 <sup>3</sup> |
| 0 | 2,153 (46.3%) | 415 (34.9%) | 2,568 (44.0%) |  |
| 0.5 | 1,674 (36.0%) | 369 (31.0%) | 2,043 (35.0%) |  |
| 1 | 620 (13.3%) | 267 (22.5%) | 887 (15.2%) |  |
| 2 | 184 (4.0%) | 129 (10.8%) | 313 (5.4%) |  |
| 3 | 15 (0.3%) | 9 (0.8%) | 24 (0.4%) |  |
| Global spoken lexical retrieval (GSLR)<sup>4</sup> | -0.00 (1.00) | -0.15 (1.10) | -0.03 (1.02) | <0.001 |
| Animals | 11.2 (5.2) | 10.7 (5.5) | 11.1 (5.2) | 0.002 |
| Vegetables | 6.8 (3.7) | 6.3 (3.9) | 6.7 (3.8) | <0.001 |
| Category fluency<sup>4</sup> | 0.00 (1.00) | -0.13 (1.09) | -0.03 (1.02) | <0.001 |
| F fluency | 9.9 (4.9) | 8.8 (5.1) | 9.7 (4.9) | <0.001 |
| L fluency | 9.2 (4.8) | 8.2 (4.9) | 9.0 (4.8) | <0.001 |
| Letter fluency<sup>4</sup> | -0.00 (1.00) | -0.24 (1.08) | -0.05 (1.02) | <0.001 |
| Multilingual Naming Test (MINT) | 24.2 (6.9) | 24.3 (6.9) | 24.2 (6.9) | 0.396 |
| Montreal Cognitive Assessment (MoCA) |  |  |  |  |
| Naming | 2.3 (0.9) | 2.4 (0.9) | 2.4 (0.9) | <0.001 |
| Naming<sup>4</sup> | 0.00 (1.00) | 0.11 (1.01) | 0.02 (1.00) | <0.001 |

1. Wilcoxon Rank Sum tests for continuous variables, Fisher's exact test for categorical.
2. P-value is 0.001 for White vs all others.
3. P-value is for categories 2 and 3 combined.
4. Standardized to mean 0, Standard deviation (SD) 1 in first AD visit age 65 or later.

**Table 2.** Language items in NACC UDS3-NB.

| Item Name | Item Description | Domain Subtype |
| --- | --- | --- |
| animals | Tell me all the animals you can think of in one minute | Category fluency |
| vegetables | Tell me all the vegetables you can think of in one minute | Category fluency |
| udsverfc | Number of correct F-words generated in 1 minute | Letter fluency |
| udsverlc | Number of correct L-words generated in 1 minute | Letter fluency |
| minttots | Multilingual naming test - total score | Naming |
| mocanami | MoCA - naming (lion, rhino, camel) | Naming |
| mocarepe | MoCA - repetition | Repetition |

### 2.3 Domain subtype-specific scores

We constructed three subdomain specific scores; a) category fluency (animals and vegetables); b) letter fluency (F-fluency, L-fluency); and c) naming (Multilingual Naming Test (MINT) [39] and MoCA [28] naming tasks). We excluded repetition from this step in the analyses as there was a single MoCA item for this secondary domain. We employed a weighted average approach using standardized item responses. Prior to score computation, all items were standardized to ensure comparability across measures. The weights applied in the averaging process were derived from the standardized factor loadings obtained through confirmatory factor analysis (CFA) models calibrated on our item bank. This method allowed us to account for the relative contribution of each item to the underlying construct, thereby producing subdomain scores that more accurately reflect the latent language abilities being assessed (see Table 3).

**Table 3.** Range and Standardized Loadings for Language Subdomain Items.

| UDS3 Items by Subdomain | Original Range | Standardized Factor Loading |
| --- | --- | --- |
| <u>Category fluency</u> |  |  |
| Animals | 0-54 | 0.895 |
| Vegetables | 0-60 | 0.879 |
| <u>Letter fluency</u> |  |  |
| F | 0-40 | 0.539 |
| L | 0-40 | 0.714 |
| <u>Naming</u> |  |  |
| MINT total score | 0-32 | 0.788 |
| MoCA Naming | 0-3 | 0.74 |

### 2.4 Statistical methods

Univariate comparisons of EOAD and LOAD were tested with Wilcoxon Rank Sum tests for continuous outcomes and Fisher’s exact test for categorical outcomes. To describe the effect of age of onset, CDR® sum of boxes (rated from 0-18 on a continuous scale, summing performance across the six domains evaluated in the global CDR®), demographics, and *APOE* genotype (coded as ≥1 ε4 allele vs. 0 ε4 alleles) on language scores, we used linear regression, with robust standard errors because the residuals in the naming model were skewed. We were also interested in how a combination of the CDR® language [40] score (clinician-rated language ability based on a combination of informal and standardized assessment, scored on the same scale as the global CDR® [37]), our subdomain scores for category fluency, letter fluency, naming, and global spoken lexical retrieval (GSLR) characterized EOAD and LOAD. We used modified Poisson regressions [41], adjusting for the CDR® sum of boxes, sex, education, race, and the presence of any *APOE* ε4 alleles. Regression assumptions were tenable in all models. We conducted sensitivity analyses using the same procedures by restricting the sample to participants biomarker-confirmed AD, defined as having both abnormally low amyloid and abnormally elevated Tau or pTau in the CSF or abnormally elevated amyloid on PET (EOAD = 401, LOAD = 738).

## 3.0 RESULTS

There were univariate differences between EOAD and LOAD for all the demographic characteristics, *APOE* ε4 genotype, and all the language measures except the MINT (Table 1). Education was fairly evenly matched; over 75% of both groups received a college degree or graduate education (see Table 1). The majority of people with LOAD and people with EOAD were white, with somewhat more racial diversity in the LOAD sample. More participants with EOAD had ≥1 *APOE* ε4 alleles. At their first NACC visit with AD dementia, 34% of the participants with EOAD received scores of mild to severe impairment (1-3) for CDR® language, almost double that of people with LOAD (18%; see Figure 1). For both groups, close to a third of scores corresponded to questionable impairment on the CDR® scale (0.5); this was slightly higher for people with LOAD. Close to half of the people with LOAD had normal language performance (CDR® language = 0).

**Figure 1.**
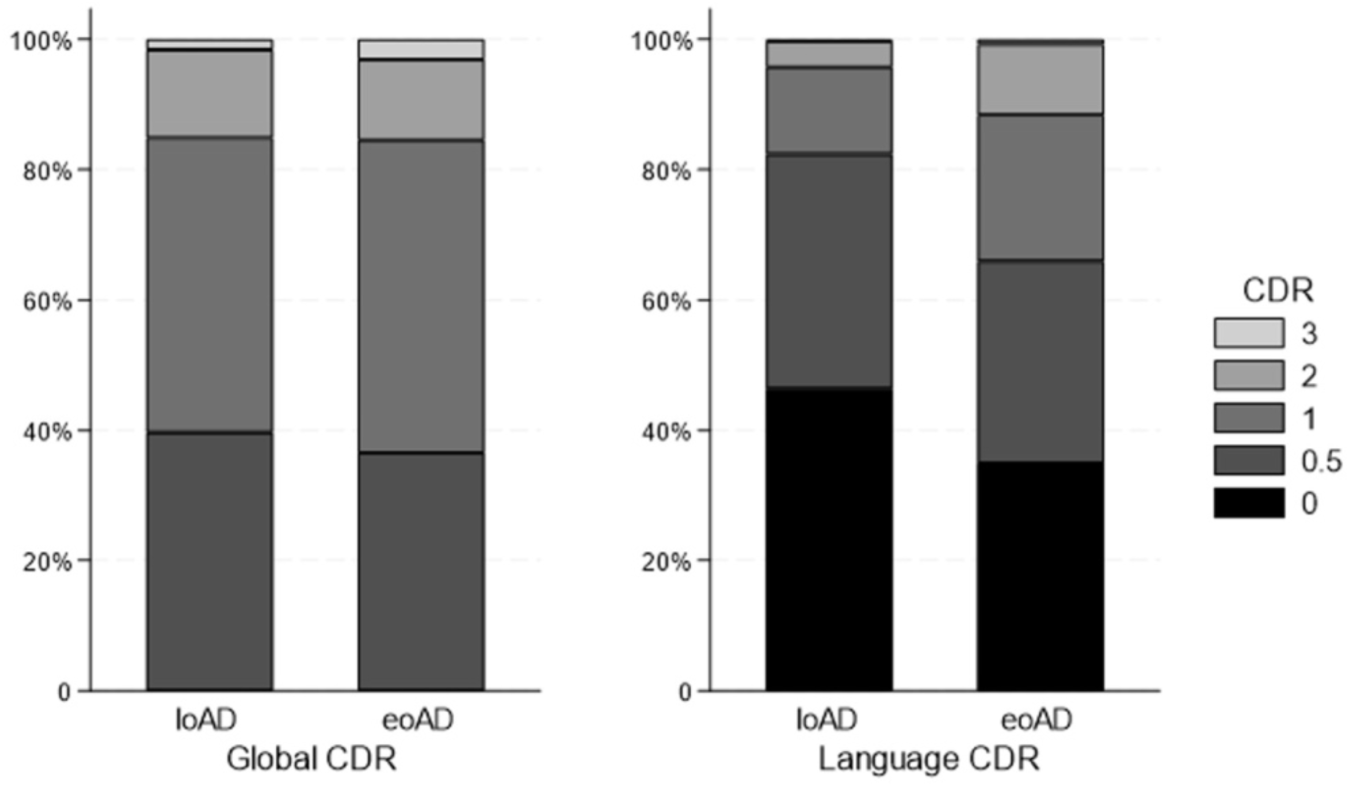
Global and Language CDR® scores in LOAD and EOAD.

Linear regression models for our language scores found statistically significantly lower GSLR, category fluency and letter fluency scores, and higher naming scores for participants with EOAD (Table 4a), despite there being a non-significant difference in CDR® sum of boxes between groups. The largest difference was observed for letter fluency, where those with EOAD were nearly a quarter SD lower. The models in Table 4b also include the effect of the CDR® language scores. Higher CDR® language scores (e.g., worse performance) were associated with lower scores (also worse performance) in GSLR, category fluency, and letter fluency in all models. In addition, when including CDR® in the regression models, the differences between EOAD and LOAD were attenuated for GSLR, category fluency, and letter fluency; only letter fluency remained statistically significant. The difference in naming was stronger when including the CDR® in the regression models, nearly a quarter of a SD higher in EOAD.

**Table 4a.**
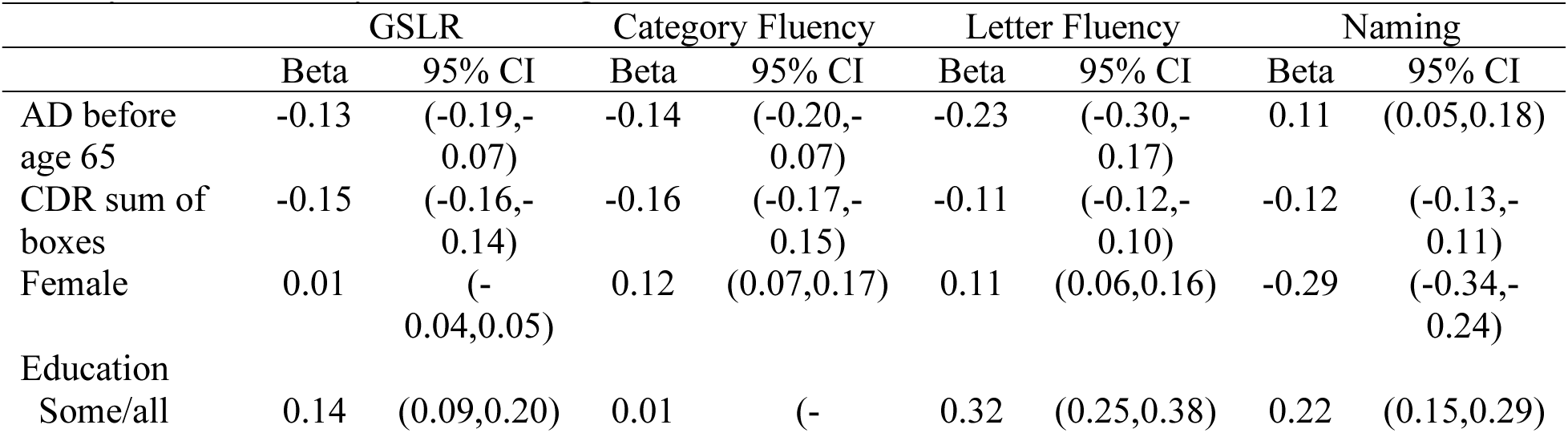
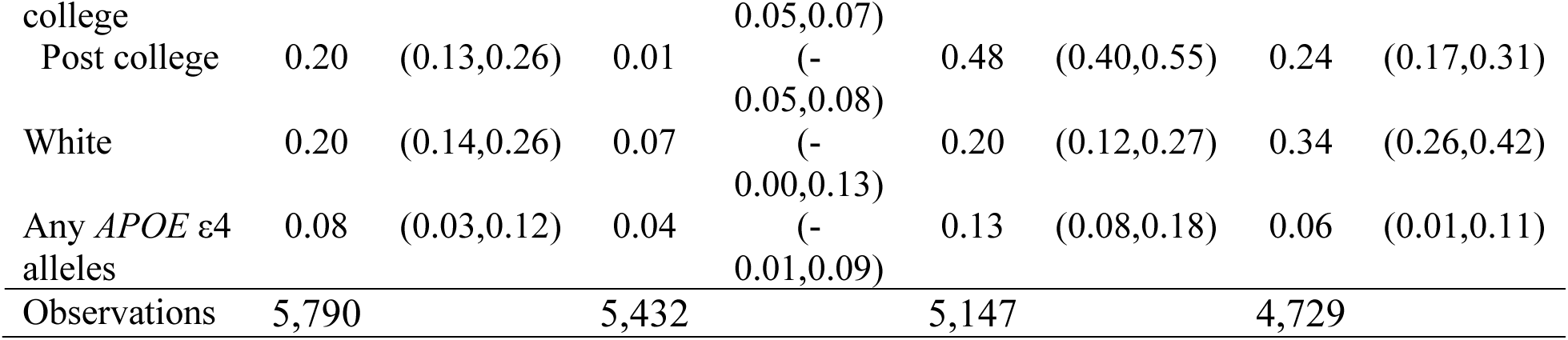
Linear regression models for global spoken lexical retrieval (GSLR), category fluency, letter fluency, and naming.

**Table 4b.** Linear regression models for global spoken lexical retrieval (GSLR), category fluency, letter fluency, and naming, additionally including the CDR language score.

|  | GSLR |  | Category Fluency |  | Letter Fluency |  | Naming |  |
| --- | --- | --- | --- | --- | --- | --- | --- | --- |
|  | Beta | 95% CI | Beta | 95% CI | Beta | 95% CI | Beta | 95% CI |
| AD before age 65 | 0.02 | (- 0.04,0.07) | -0.01 | (- 0.07,0.05) | -0.14 | (-0.20,- 0.07) | 0.24 | (0.18,0.30) |
| CDR sum of boxes | -0.10 | (-0.11,- 0.09) | -0.12 | (-0.12,- 0.11) | -0.08 | (-0.09,- 0.07) | -0.08 | (-0.09,- 0.07) |
| CDR language |  |  |  |  |  |  |  |  |
| 0.5 | -0.27 | (-0.32,- 0.23) | -0.25 | (-0.30,- 0.20) | -0.15 | (-0.21,- 0.09) | -0.18 | (-0.23,- 0.13) |
| 1 | -0.81 | (-0.88,- 0.75) | -0.72 | (-0.80,- 0.65) | -0.49 | (-0.57,- 0.41) | -0.70 | (-0.79,- 0.61) |
| 2 or 3 | -1.27 | (-1.39,- 1.15) | -1.26 | (-1.39,- 1.13) | -1.10 | (-1.24,- 0.96) | -1.30 | (-1.48,- 1.11) |
| Female | -0.03 | (- 0.07,0.02) | 0.09 | (0.05,0.14) | 0.09 | (0.04,0.14) | -0.32 | (-0.37,- 0.27) |
| Education |  |  |  |  |  |  |  |  |
| Some/all college | 0.15 | (0.10,0.20) | 0.01 | (- 0.04,0.07) | 0.32 | (0.25,0.38) | 0.23 | (0.17,0.30) |
| Post college | 0.22 | (0.16,0.27) | 0.03 | (- 0.03,0.10) | 0.48 | (0.42,0.55) | 0.27 | (0.20,0.34) |
| Race (% White) | 0.24 | (0.18,0.29) | 0.10 | (0.03,0.16) | 0.22 | (0.15,0.29) | 0.36 | (0.28,0.44) |
| Any *APOE* ε4 alleles | 0.05 | (0.01,0.10) | 0.02 | (- 0.03,0.06) | 0.11 | (0.06,0.16) | 0.03 | (- 0.02,0.08) |
| Observations | 5,790 |  | 5,432 |  | 5,147 |  | 4,729 |  |

We also examined which combination of CDR® language ratings and domain-specific and global language scores best differentiated average abilities for people with EOAD compared to people with LOAD. Compared to those with LOAD, individuals with EOAD had worse CDR® language and letter fluency scores, but better naming scores (see Table 5). Of note, these differences were also observed when we controlled for overall dementia severity by including a term for the global CDR® rating. Further, we also included a term for the CDR® sum of boxes, which captures aspects of disease progression in a more granular way than the overall global score.

**Table 5.**
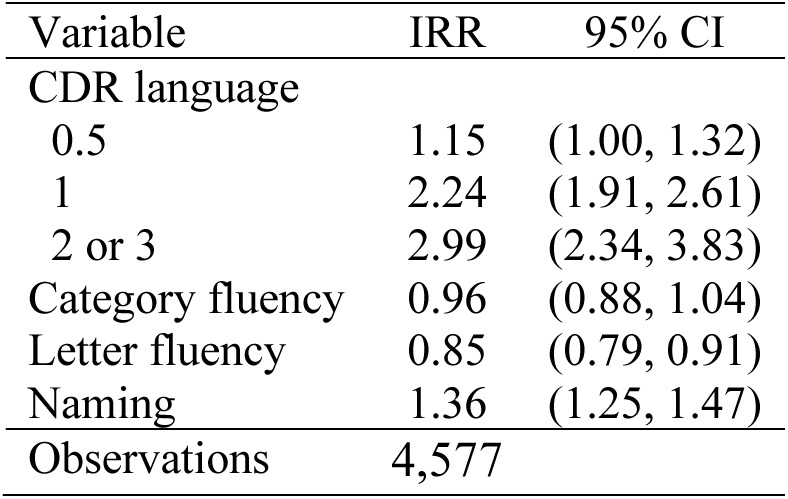
Modified Poisson regression for AD before age 65 (Incidence rate ratio, IRR, and 95% CI), adjusted for CDR sum of boxes, sex, education, race, and any *APOE* ε4 alleles.

| Variable | IRR | 95% CI |
| --- | --- | --- |
| CDR language |  |  |
| 0.5 | 1.15 | (1.00, 1.32) |
| 1 | 2.24 | (1.91, 2.61) |
| 2 or 3 | 2.99 | (2.34, 3.83) |
| Category fluency | 0.96 | (0.88, 1.04) |
| Letter fluency | 0.85 | (0.79, 0.91) |
| Naming | 1.36 | (1.25, 1.47) |
| Observations | 4,577 |  |

Sensitivity analyses restricted to people with biomarker-confirmed AD generally confirmed the primary analyses. Without adjustment for CDR® language, differences were attenuated for GLSR and category fluency (Supplementary Table 1a) but of similar magnitude, when adjusted for CDR® language (Supplementary Table 1b). In the examination of the relative effects of the language measures, most estimates were similar; only the CDR® language effects were reduced (Supplementary Table 1c).

## 4.0 DISCUSSION

This study examined the characteristics of several aspects of language including spoken language in people with EOAD and LOAD using standardized assessments of lexical retrieval and more global spoken language ability through several specific tests as well as the CDR® derived from interviews with informants and provider ratings. We found that participants with LOAD had higher language scores on average than people with EOAD for category fluency, letter fluency, and for global spoken lexical retrieval. In contrast, we did not see large differences on average between people with EOAD and people with LOAD in terms of confrontation naming ability.

Several previous studies have shown differences in speech, language, and communication in early-onset dementias, characterizing rarer forms of language-led syndromes (e.g., the logopenic variant of primary progressive aphasia) as early-onset rather than late-onset. Notably, prior detailed investigations of cognitive subdomains including language in people with EOAD and people with LOAD are of a smaller scale and restricted to single study sites [6]. Hammers *et al.* [6] provided a larger multi-site comparison of cognitive profiles in EOAD and LOAD through the Alzheimer’s Disease Neuroimaging Initiative (ADNI [42]) and Longitudinal Early-Onset Alzheimer’s Disease Study cohorts (LEADS [43]). Their findings suggest that, despite previous findings indicating worse performance in non-amnestic domains in EOAD, language may be more impaired in LOAD, with substantial variability in both groups. Based on the averaged raw scores alone, people with EOAD had worse scores on average than those with LOAD for category fluency, had similar scores on average compared to those with LOAD for confrontation naming and story recall, and had better scores on average compared to those with LOAD for word recognition. For the composite language score, however, people with LOAD had lower scores on average than people with EOAD overall. In contrast, an independent meta-analysis of 42 studies on the cognitive profiles of EOAD and LOAD by Seath *et al.* [44] found that people with EOAD differ significantly from people with LOAD at baseline: a greater proportion of participants with early-onset dementia exhibited atypical presentations with additional impairments to non-amnestic domains such as language and a greater overall severity of symptoms. While data from 21,856 patients were synthesized for Seath *et al.*’s [44] work (EOAD = 5,544), all cognitive domains discussed in that paper were evaluated using the MMSE. Of note, while six MMSE items assess language directly, only one (confrontation naming) specifically addresses spoken language.

Nuanced characterization and documentation of “typical” language performance in EOAD and LOAD has direct clinical implications, beyond diagnostic differentiation. Targeted behavioral intervention is paramount to enhance, maintain, or compensate for communication in people living with AD dementia, particularly for spoken language [29–31]. Speech and language therapy is the primary intervention that can slow down the behavioral impact of AD on communication [32–34] and is documented to effectively enhance life participation and well-being for people living with dementia [30,31,33,35].

Our analyses build on prior work and examine multiple features of language including spoken lexical retrieval to further evaluate symptomatic features of EOAD and LOAD. Our results suggest that profiles of language on average in people with EOAD and people with LOAD are distinct. Our results support that, on average, people with EOAD present with more severe changes in language performance at the time of dementia diagnosis and greater heterogeneity in subdomains of spoken lexical retrieval than people with LOAD. Furthermore, our work demonstrates nuanced profiling of facets of language performance. Within spoken lexical retrieval, confrontation naming alone was higher in EOAD than in LOAD. However, spoken lexical retrieval was lower on average among people with EOAD than people with LOAD for both category and letter fluency. Additionally, our GLSR scores and the CDR® language scores were lower on average in people with EOAD than people with LOAD. Our work thus suggests that tests of naming alone are insufficient to capture the differing linguistic profiles of EOAD and LOAD. Considering language as a monolithic construct, or even considering the subdomain of spoken lexical retrieval as a monolithic construct, may obscure important differences between people with EOAD and people with LOAD.

The CDR® language score also differed on average between people with EOAD and people with LOAD. The CDR® language score is ascertained and reported by specialist clinicians and is based on patient and care partner report, observation, and clinical evaluation [40]. The CDR® language score represents performance in discourse, speech mechanics, auditory comprehension, repetition, semantics, reading, and writing. We leveraged the global CDR® to further classify phenotypic presentation. While the time of diagnosis is a proxy for identifying substantial changes in cognition, the time between symptom onset and diagnosis is highly variable [45]. By examining individuals at a broadly comparable point in global clinical progression, we reduced variability in disease severity and progression that might otherwise confound comparisons between people with EOAD and people with LOAD. Furthermore, controlling for overall CDR® sum of boxes allowed us to evaluate the specific role language may play in comparison to overall global cognition. This focus helped isolate differences attributable specifically to age of onset (i.e., EOAD vs. LOAD), rather than disease stage.

Our study leveraged a uniquely robust sample of individuals with EOAD and LOAD from a large, multi-site cohort of incident and prevalent AD from NACC. The standardized data collection across Alzheimer’s Disease Research Centers (ADRCs), conducted by highly trained clinicians and research personnel with deep expertise in dementia diagnosis and assessment, ensures the clinical rigor of EOAD and LOAD classification, and ascertainment of language performance in standardized fashion across all participant encounters.This methodological rigor allows us to move beyond the limitations of single-clinician or single-site case series, providing a much broader and more reliable foundation for examining language subdomains across AD onset types.

Our study protocol in NACC allows for a more comprehensive evaluation of spoken language, with multiple items per subdomain. One limitation of this work is the lack of participant data prior to NACC enrollment: it is possible that some participants were misclassified as LOAD if they were diagnosed with AD elsewhere before age 65 but didn’t join NACC until after age 65. There was likely participant overlap with the LEADS and ADNI cohorts described in Hammers *et al.* [6]. People who participated in LEADS and ADNI agree to more research procedures and those samples may be even less generalizable than those in the NACC data set.

Our work shows that a substantial proportion of individuals with AD dementia, regardless of age of onset, exhibit decline in spoken lexical retrieval function at the time of diagnosis. Furthermore, we show that this decline is more pronounced and varied on average in people with EOAD than in people with LOAD. These findings are centered on assessments of spoken lexical retrieval: confrontation naming and verbal fluency, essential skills for daily communication. While confrontation naming targets precision and verbal fluency evaluates appropriateness and efficiency, both skills require the individual to access, retrieve, and verbally produce target lexical items—cornerstones of communicating effectively.

We found significant variability within these skills within and across groups, particularly for EOAD. Our findings support that EOAD differs from LOAD by more than age of onset and instead represents a distinct language phenotypical profile on average with more prominent and heterogeneous language deficits. This work sheds light on how AD dementia may affect communication differentially for people with EOAD and people with LOAD and may inform early detection strategies and intervention planning—particularly for younger individuals who are more likely to experience disruptions to occupation, finances, and familial dependents because of the early-onset of the condition. The present findings demonstrate that reduced language performance was common at the time of AD diagnosis in both groups; while this was somewhat more common in people with EOAD, it was not at all rare for people with LOAD. These results support that a focus on language in addition to focus on memory may be very useful to characterize these populations. Furthermore, these results suggest the possibility that many people with either EOAD or LOAD may experience clinically relevant issues with spoken communication, even at the time of AD diagnosis. Further research on the effectiveness of interventions that target improving communication in these populations is warranted.

## Data Availability

All data produced in the present study are available upon reasonable request to the authors

## 5.0 ACKNOWLEDGMENTS

This work was supported by the National Institute on Aging (NIA P50 AG005136; Shubhabrata Mukherjee: R01 AG082730; U24 AG074855 to Timothy Hohman) and the University of Washington Alzheimer’s Disease Research Center Development Project Award funded to Jeanne Gallée (NIA P30AG066509).

## 7.0 CONSENT STATEMENT

All human subjects involved in the present study provided written informed consent.

---

**Supplementary Figure 1.**
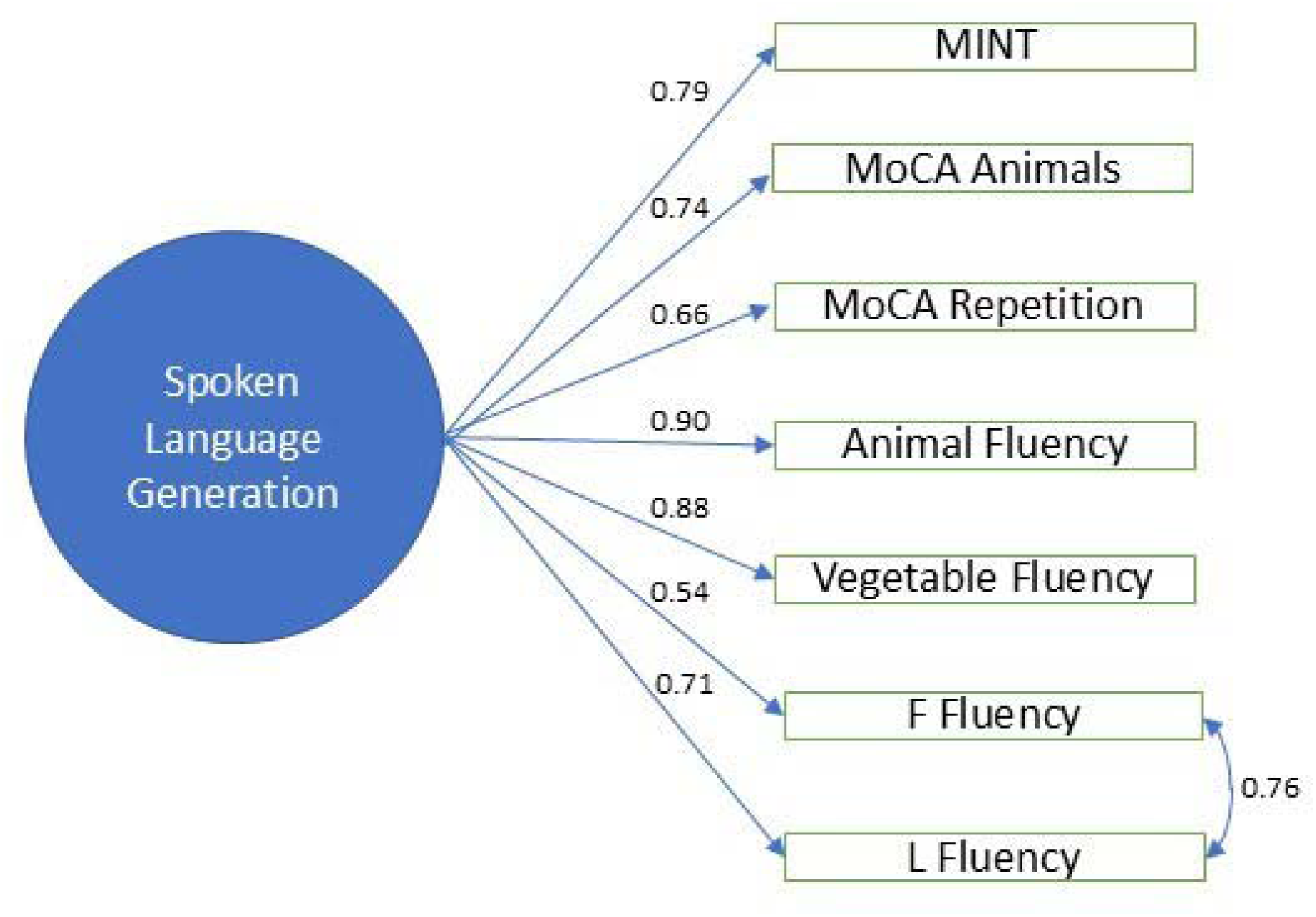
Standardized primary factor loadings for the global spoken lexical retrieval (GSLR) score, with the residual correlation between the phonemic fluency items.

## REFERENCES

1. Alzheimer’s Association. 2025 Alzheimer’s disease facts and figures. Alzheimers Dement. 2025;21(4):e12345.

2. American Psychiatric Association. Diagnostic and statistical manual of mental disorders. 5th ed. Arlington, VA: American Psychiatric Association; 2013.

3. Dubois B, Feldman HH, Jacova C, Hampel H, Molinuevo JL, Blennow K, et al. Advancing research diagnostic criteria for Alzheimer’s disease: the IWG-2 criteria. Lancet Neurol. 2014;13(6):614–29.

4. McKhann GM, Knopman DS, Chertkow H, Hyman BT, Jack CR Jr, Kawas CH, et al. The diagnosis of dementia due to Alzheimer’s disease: recommendations from the National Institute on Aging–Alzheimer’s Association workgroups on diagnostic guidelines for Alzheimer’s disease. Alzheimers Dement. 2011;7(3):263–9.

5. Mendez MF. Early-onset Alzheimer’s disease: nonamnestic subtypes and type 2 AD. Arch Med Res. 2012;43(8):677–85.

6. Hammers DB, Eloyan A, Thangarajah M, et al. Differences in baseline cognitive performance between participants with early-onset and late-onset Alzheimer’s disease: Comparison of LEADS and ADNI. Alzheimers Dement. 2025;21(1):e14218.

7. Crane PK, Trittschuh E, Mukherjee S, et al. Incidence of cognitively defined late-onset Alzheimer’s dementia subgroups from a prospective cohort study. Alzheimers Dement. 2017;13(12):1307–16.

8. Mukherjee S, Mez J, Trittschuh EH, et al. Genetic data and cognitively defined late-onset Alzheimer’s disease subgroups. Mol Psychiatry. 2020;25(11):2942–51.

9. Crane PK, Groot C, Ossenkoppele R, et al. Cognitively defined Alzheimer’s dementia subgroups have distinct atrophy patterns. Alzheimers Dement. 2024;20(3):1739–52.

10. Kim J, Woo SY, Kim S, et al. Differential effects of risk factors on the cognitive trajectory of early- and late-onset Alzheimer’s disease. Alzheimers Res Ther. 2021;13(1):113.

11. Tort-Merino A, Falgàs N, Allen IE, et al. Early-onset Alzheimer’s disease shows a distinct neuropsychological profile and more aggressive trajectories of cognitive decline than late-onset. Ann Clin Transl Neurol. 2022;9(12):1962–73.

12. Vonk JM, Rentería MA, Geerlings MI, Avila JF, Mayeux R, Manly JJ. When verbal fluency inverts: Temporality of semantic impairment in preclinical Alzheimer’s disease. Alzheimers Dement. 2021;17(S6):e053877.

13. Kaltsa M, Tsolaki A, Lazarou I, et al. Language Markers of Dementia and Their Role in Early Diagnosis of Alzheimer’s Disease: Exploring Grammatical and Syntactic Competence via Sentence Repetition. J Alzheimers Dis Rep. 8(1):1115–32.

14. Klimova B, Kuca K. Speech and language impairments in dementia. Journal of Applied Biomedicine. 2016 Apr 1;14(2):97–103.

15. Yeung A, Iaboni A, Rochon E, et al. Correlating natural language processing and automated speech analysis with clinician assessment to quantify speech-language changes in mild cognitive impairment and Alzheimer’s dementia. Alzheimers Res Ther. 2021;13(1):109.

16. Forbes-McKay K, Shanks MF, Venneri A. Profiling spontaneous speech decline in Alzheimer’s disease: a longitudinal study. Acta Neuropsychiatr. 2013;25(6):320–7.

17. Mueller KD, Koscik RL, LaRue A, Clark LR, Hermann B, Johnson SC, et al. Verbal Fluency and Early Memory Decline: Results from the Wisconsin Registry for Alzheimer’s Prevention. Archives of Clinical Neuropsychology. 2015 Aug 1;30(5):448– 57.

18. Palasí A, Gutiérrez-Iglesias B, Alegret M, et al. Differentiated clinical presentation of early and late-onset Alzheimer’s disease: is 65 years of age providing a reliable threshold? J Neurol. 2015;262(5):1238–46.

19. Imamura T, Takatsuki Y, Fujimori M, et al. Age at onset and language disturbances in Alzheimer’s disease. Neuropsychologia. 1998;36(9):945–9.

20. Tellechea P, Pujol N, Esteve-Belloch P, et al. Early- and late-onset Alzheimer disease: Are they the same entity? Neurología (Engl Ed). 2018;33(4):244–53.

21. Kaiser NC, Melrose RJ, Liu C, Sultzer DL, Jimenez E, Su M, et al. Neuropsychological and neuroimaging markers in early versus late-onset Alzheimer’s disease. Am J Alzheimers Dis Other Demen. 2012 Nov;27(7):520–9.

22. Koss E, Edland S, Fillenbaum G, Mohs R, Clark C, Galasko D, et al. Clinical and neuropsychological differences between patients with earlier and later onset of Alzheimer’s disease: A CERAD analysis, Part XII. Neurology. 1996 Jan;46(1):136–41.

23. Joubert S, Gour N, Guedj E, Didic M, Guériot C, Koric L, et al. Early-onset and late-onset Alzheimer’s disease are associated with distinct patterns of memory impairment. Cortex. 2016 Jan 1;74:217–32.

24. Suribhatla S, Baillon S, Dennis M, Marudkar M, Muhammad S, Munro D, et al. Neuropsychological performance in early and late onset Alzheimer’s disease: comparisons in a memory clinic population. International Journal of Geriatric Psychiatry. 2004;19(12):1140–7.

25. Licht EA, McMurtray AM, Saul RE, Mendez MF. Cognitive differences between early- and late-onset Alzheimer’s disease. Am J Alzheimers Dis Other Demen. 2007;22(3):218–22.

26. Grady CL, Haxby JV, Horwitz B, Berg G, Rapoport SI. Neuropsychological and cerebral metabolic function in early vs late onset dementia of the Alzheimer type. Neuropsychologia. 1987 Jan 1;25(5):807–16.

27. Folstein MF, Folstein SE, McHugh PR. “Mini-mental state”. A practical method for grading the cognitive state of patients for the clinician. J Psychiatr Res. 1975 Nov;12(3):189–98.

28. Nasreddine ZS, Phillips NA, Bédirian V, et al. The Montreal Cognitive Assessment, MoCA: a brief screening tool for mild cognitive impairment. J Am Geriatr Soc. 2005;53(4):695–9.

29. Bayles KA. Understanding the Neuropsychological Syndrome of Dementia. Semin Speech Lang. 2001;22(04):251–60.

30. Volkmer A, Cross L, Highton L, et al. ’Communication is difficult’: Speech, language and communication needs of people with young onset or rarer forms of non-language led dementia. Int J Lang Commun Disord. 2024;59(4):1553–77.

31. Volkmer A, editor. Assessment and therapy for language and cognitive communication difficulties in dementia and other progressive diseases. 2nd ed. J & R Press; 2024.

32. American Speech-Language-Hearing Association. Scope of Practice in Speech-Language Pathology [Internet]. 2016 [cited 2025 May 25]. Available from: https://www.asha.org/policy/sp2016-00343/

33. Hoffman P. Assessment and therapy for language and communication difficulties in dementia and other progressive diseases. Neuropsychol Rehabil. 2014;24(2):302–3.

34. Swan K, Hopper M, Wenke R, Jackson C, Till T, Conway E. Speech-Language Pathologist Interventions for Communication in Moderate–Severe Dementia: A Systematic Review. Am J Speech Lang Pathol. 2018;27(2):836–52.

35. Hickey & Bourgeois, 2009

36. Weintraub S, Besser L, Dodge HH, et al. Version 3 of the Alzheimer disease centers’ neuropsychological test battery in the Uniform Data Set (UDS). Alzheimer Dis Assoc Disord. 2018;32(1):10–17. doi: 10.1097/WAD.0000000000000223

37. Morris JC, Ernesto C, Schafer K, Coats M, Leon S, Sano M, et al. Clinical dementia rating training and reliability in multicenter studies: the Alzheimer’s Disease Cooperative Study experience. Neurology. 1997 Jun;48(6):1508–10.

38. Mukherjee S, Choi SE, Lee ML, et al. Cognitive domain harmonization and cocalibration in studies of older adults. Neuropsychology. 2023;37(4):409–23.

39. Gollan TH, Weissberger GH, Runnqvist E, Montoya RI, Cera CM. Self-ratings of Spoken Language Dominance: A Multi-Lingual Naming Test (MINT) and Preliminary Norms for Young and Aging Spanish-English Bilinguals. Biling (Camb Engl). 2012;15(3):594–615.

40. Knopman DS, Weintraub S, Pankratz VS. Language and behavior domains enhance the value of the clinical dementia rating scale. Alzheimers Dement. 2011;7(3):293–9.

41. Zou G. A modified poisson regression approach to prospective studies with binary data. American journal of epidemiology. 2004 Apr 1;159(7):702–6.

42. ADNI3. Alzheimer’s Disease Neuroimaging Initiative: ADNI3 Procedures Manual [Internet]. 2021 [cited 2025 May 25]. Available from: https://adni.loni.usc.edu/wp-content/upLOADs/2012/10/ADNI3-Procedures-Manual_v3.0_20170627.pdf

43. Apostolova LG, Aisen P, Eloyan A, et al. The Longitudinal Early-onset Alzheimer’s Disease Study (LEADS): Framework and methodology. Alzheimers Dement. 2021;17(12):2043–55.

44. Seath P, Macedo-Orrego LE, Velayudhan L. Clinical characteristics of early-onset versus late-onset Alzheimer’s disease: a systematic review and meta-analysis. Int Psychogeriatr. 2024;36(12):1093–109.

45. Kerwin D, Abdelnour C, Caramelli P, et al. Alzheimer’s disease diagnosis and management: Perspectives from around the world. Alzheimers Dement (Amst). 2022;14(1):e12334.

