## Supplementary Table 1a for "Facets of language performance in early-onset and late-onset Alzheimer’s disease dementia"

**Table 1a. Linear regression models for global spoken lexical retrieval (GSLR), category fluency, letter fluency, and naming.**

The differences between EOAD and LOAD were smaller than in the full sample for GSLR ($\beta$ = -0.08; 95% CI (-0.19,0.04)) and catflu ($\beta$ = -0.10; 95% CI (-0.21,0.02)).

| | GSLR | | Category Fluency | | Letter Fluency | | Naming | |
|---|---|---|---|---|---|---|---|---|
| | Beta | 95% CI | Beta | 95% CI | Beta | 95% CI | Beta | 95% CI |
| AD before age 65 | -0.08 | (-0.19,0.04) | -0.10 | (-0.21,0.02) | -0.24 | (-0.37,-0.11) | 0.15 | (0.02,0.27) |
| CDR sum of boxes | -0.16 | (-0.18,-0.14) | -0.17 | (-0.19,-0.15) | -0.13 | (-0.15,-0.10) | -0.11 | (-0.13,-0.08) |
| Female | 0.05 | (-0.06,0.16) | 0.20 | (0.09,0.31) | 0.10 | (-0.02,0.22) | -0.35 | (-0.47,-0.24) |
| Education | | | | | | | | |
| Some/all college | 0.13 | (-0.01,0.27) | 0.07 | (-0.08,0.22) | 0.28 | (0.13,0.43) | 0.21 | (0.04,0.37) |
| Post college | 0.17 | (0.01,0.33) | 0.09 | (-0.07,0.25) | 0.51 | (0.35,0.67) | 0.11 | (-0.07,0.29) |
| White | 0.11 | (-0.07,0.30) | -0.01 | (-0.21,0.19) | 0.12 | (-0.10,0.34) | 0.24 | (-0.00,0.48) |
| Any *APOE* ε4 alleles | 0.08 | (-0.03,0.19) | 0.03 | (-0.08,0.14) | 0.06 | (-0.06,0.18) | 0.06 | (-0.05,0.18) |
| Observations | 1,133 | | 1,056 | | 1,007 | | 963 | |
