## Supplementary Table 1b for "Facets of language performance in early-onset and late-onset Alzheimer’s disease dementia"

**Table 1b. Linear regression models for global spoken lexical retrieval (GSLR), category fluency, letter fluency, and naming, additionally including the CDR language score.** The EOAD differences are similar to those in the full sample.

| | GSLR | | Category Fluency | | Letter Fluency | | Naming | |
|---|---|---|---|---|---|---|---|---|
| | Beta | 95% CI | Beta | 95% CI | Beta | 95% CI | Beta | 95% CI |
| AD before age 65 | -0.00 | (-0.11,0.10) | -0.03 | (-0.14,0.08) | -0.18 | (-0.30,-0.05) | 0.22 | (0.10,0.34) |
| CDR sum of boxes | -0.11 | (-0.13,-0.08) | -0.12 | (-0.14,-0.10) | -0.09 | (-0.11,-0.06) | -0.07 | (-0.10,-0.04) |
| CDR language | | | | | | | | |
| 0.5 | -0.28 | (-0.40,-0.17) | -0.27 | (-0.38,-0.15) | -0.15 | (-0.28,-0.01) | -0.12 | (-0.24,0.00) |
| 1 | -0.88 | (-1.04,-0.73) | -0.72 | (-0.89,-0.56) | -0.59 | (-0.76,-0.41) | -0.65 | (-0.83,-0.47) |
| 2 or 3 | -1.29 | (-1.58,-1.00) | -1.24 | (-1.55,-0.93) | -1.15 | (-1.47,-0.82) | -1.12 | (-1.50,-0.75) |
| Female | 0.01 | (-0.09,0.11) | 0.16 | (0.06,0.27) | 0.07 | (-0.05,0.18) | -0.37 | (-0.48,-0.26) |
| Education | | | | | | | | |
| Some/all college | 0.15 | (0.01,0.28) | 0.08 | (-0.06,0.22) | 0.29 | (0.15,0.43) | 0.23 | (0.08,0.39) |
| Post college | 0.20 | (0.05,0.34) | 0.11 | (-0.04,0.26) | 0.53 | (0.37,0.69) | 0.15 | (-0.02,0.32) |
| White | 0.20 | (0.02,0.38) | 0.06 | (-0.14,0.26) | 0.17 | (-0.05,0.38) | 0.29 | (0.06,0.53) |
| Any *APOE* ε4 alleles | 0.06 | (-0.04,0.16) | 0.00 | (-0.10,0.11) | 0.04 | (-0.08,0.15) | 0.02 | (-0.09,0.14) |

| | | | | |
|---|---|---|---|---|
| Observations | 1,133 | 1,056 | 1,007 | 963 |
