## Supplementary Table 1C for "Facets of language performance in early-onset and late-onset Alzheimer’s disease dementia"

**Table 1c. Modified Poisson regression for AD before age 65 (**Incidence rate ratio, IRR, and 95% CI)**, adjusted for CDR sum of boxes, sex, education, race, and any *APOE* ε4 alleles.** The IRRs for our subscales are similar to the full sample, but CDR language effects are attenuated.

| Variable | IRR | 95% CI |
|---|---|---|
| CDR language | | |
| 0.5 | 0.87 | (0.71, 1.07) |
| 1 | 1.24 | (0.97, 1.59) |
| 2 or 3 | 1.87 | (1.31, 2.68) |
| Category fluency | 0.93 | (0.82, 1.06) |
| Letter fluency | 0.86 | (0.77, 0.95) |
| Naming | 1.28 | (1.13, 1.47) |
| Observations | 929 | |
